# Redox imbalance and oxidative DNA damage during isoniazid treatment: A clinical and translational pharmacokinetic study

**DOI:** 10.1101/2020.04.14.20065292

**Authors:** Isaac Zentner, Hyun-moon Back, Leonid Kagan, Selvakumar Subbian, Jyothi Nagajyothi, Shashikant Srivastava, Jotam Pasipanodya, Tawanda Gumbo, Gregory P. Bisson, Christopher Vinnard

**Affiliations:** Public Health Research Institute, New Jersey Medical School, Newark, NJ, USA; Ernest Mario School of Pharmacy, Rutgers University, Piscataway, NJ, USA; Jerry H. Hodge School of Pharmacy, Texas Tech University Health Sciences Center, Lubbock, TX, USA; University of Pennsylvania, Perelman School of Medicine, Philadelphia, PA, USA

## Abstract

**Background:** The potential for hepatotoxicity during isoniazid-based tuberculosis (TB) treatment presents a major challenge for TB control programs worldwide. We sought to determine whether pharmacokinetic exposures of isoniazid and its metabolites were related to cellular oxidation/reduction status and downstream markers of oxidative DNA damage.

**Methods:** We performed intensive pharmacokinetic sampling among isoniazid-treated patients to determine the relative plasma exposures of isoniazid, acetylisoniazid, hydrazine, and acetylhydrazine. Physiologically-based pharmacokinetic modeling was used to estimate liver tissue exposures during a 24-hour dosing interval for each compound. We experimentally treated HepG2 cells with isoniazid and metabolites at equimolar concentrations corresponding to these exposures for 7, 14, and 28 day periods, and performed assays related to redox imbalance and oxidative DNA damage at each timepoint. We related a urine marker of oxidative DNA damage to serum isoniazid pharmacokinetic exposures and pharmacogenetics in a clinical study.

**Results:** Among isoniazid-treated patients, serum concentrations of hydrazine and isoniazid concentrations were highly correlated. At equimolar concentrations that approximated hepatic tissue exposures during a 24-hour dosing interval, hydrazine demonstrated the highest levels of redox imbalance, mitochondrial injury, and oxidative DNA damage over a 28-day treatment period. In a clinical validation study of isoniazid-treated TB patients, peak isoniazid serum concentrations were positively associated with a urine biomarker of oxidative DNA damage.

**Conclusions:** Isoniazid and its metabolites share the potential for oxidative cellular damage, with the greatest effects observed for hydrazine. Future studies should investigate the clinical consequences of oxidative stress with regards to clinical episodes of drug induced liver injury during isoniazid treatment.

## BACKGROUND

The potential for isoniazid hepatotoxicity during tuberculosis (TB) treatment remains a major challenge for TB control efforts worldwide. In the United States, isoniazid is the second leading cause of drug-induced liver injury, with evidence of under-reporting^1^. Aside from the direct effects of patient injury, the potential for hepatotoxicity during isoniazid treatment places additional burdens on TB control programs^2^, including laboratory monitoring and treatment interruptions, and hampers drug development efforts^3^. The risk of isoniazid-related hepatotoxicity is further increased in the setting of co-infection with human immunodeficiency virus (HIV)^4^.

A central challenge to understanding isoniazid hepatotoxicity has been its idiosyncratic nature, lacking a clear relationship with dose size^5^. Damage mediated by reactive oxygen species (ROS), as a consequence of imbalance between cellular oxidation and reduction^6^, is a key determinant of drug hepatotoxicity^7,8^. The imbalance between oxidation and reduction processes overwhelms cellular stores of glutathione, which provides an intracellular sink for ROS directed by the glutathione-s- transferase (GST) family of proteins^9^. With its high cellular density of mitochondria, the liver is particularly vulnerable to injury from ROS byproducts, leading to free radical-induced modifications of host proteins, DNA, and lipids^10^.

While there is abundant evidence that isoniazid and its metabolites cause redox imbalance in mammalian cells^5,11,12^, clinical studies have not yet linked isoniazid parent-metabolite pharmacokinetic exposures with markers of oxidative stress. Hydrazine, an isoniazid metabolite that damages the electron transport chain in mitochondria, has been proposed as a primary driver of isoniazid hepatotoxicity, yet pre-clinical studies have evaluated concentrations higher than those achieved in patients during isoniazid treatment. Establishing the link, if any, between markers of oxidative stress and hepatotoxicity during isoniazid treatment could lead to alternate approaches to monitoring during treatment, personalization of isoniazid dosing, or the development of ameliorating therapeutic strategies for toxicity prevention^13^. Here, we hypothesized that long-term exposures to isoniazid and its metabolites were related to markers of redox imbalance in hepatocytes, and that hydrazine exposures would drive redox imbalance and oxidative damage in the liver.

## METHODS

### Clinical pharmacokinetic study to determine relative plasma exposures of isoniazid, acetylisoniazid, hydrazine, and acetylhydrazine

We performed intensive pharmacokinetic sampling for drug and metabolite concentrations in 10 TB patients (4 latent infection patients and 6 active TB patients) treated with isoniazid, in order to identify the relevant metabolite concentrations for subsequent pre-clinical experiments. After an overnight fast, patients were dosed with oral isoniazid (300 mg). A pre-dose blood draw was performed, with additional blood draws at 1, 2, 4, 6, and 8 hours after dosing. Following centrifugation, plasma samples were stored at -80C until shipment to Alturas Analytics (Moscow, ID) for analysis. At each timepoint, we measured plasma concentrations of isoniazid, acetylisoniazid, hydrazine, and acetylhydrazine (Figure 1). Noncompartmental pharmacokinetic analysis was performed to determine area under the concentration-versus-time curve (AUC), extrapolated over a 24-hour dosing interval according to the slope defined in the elimination phase.

**Figure 1.**
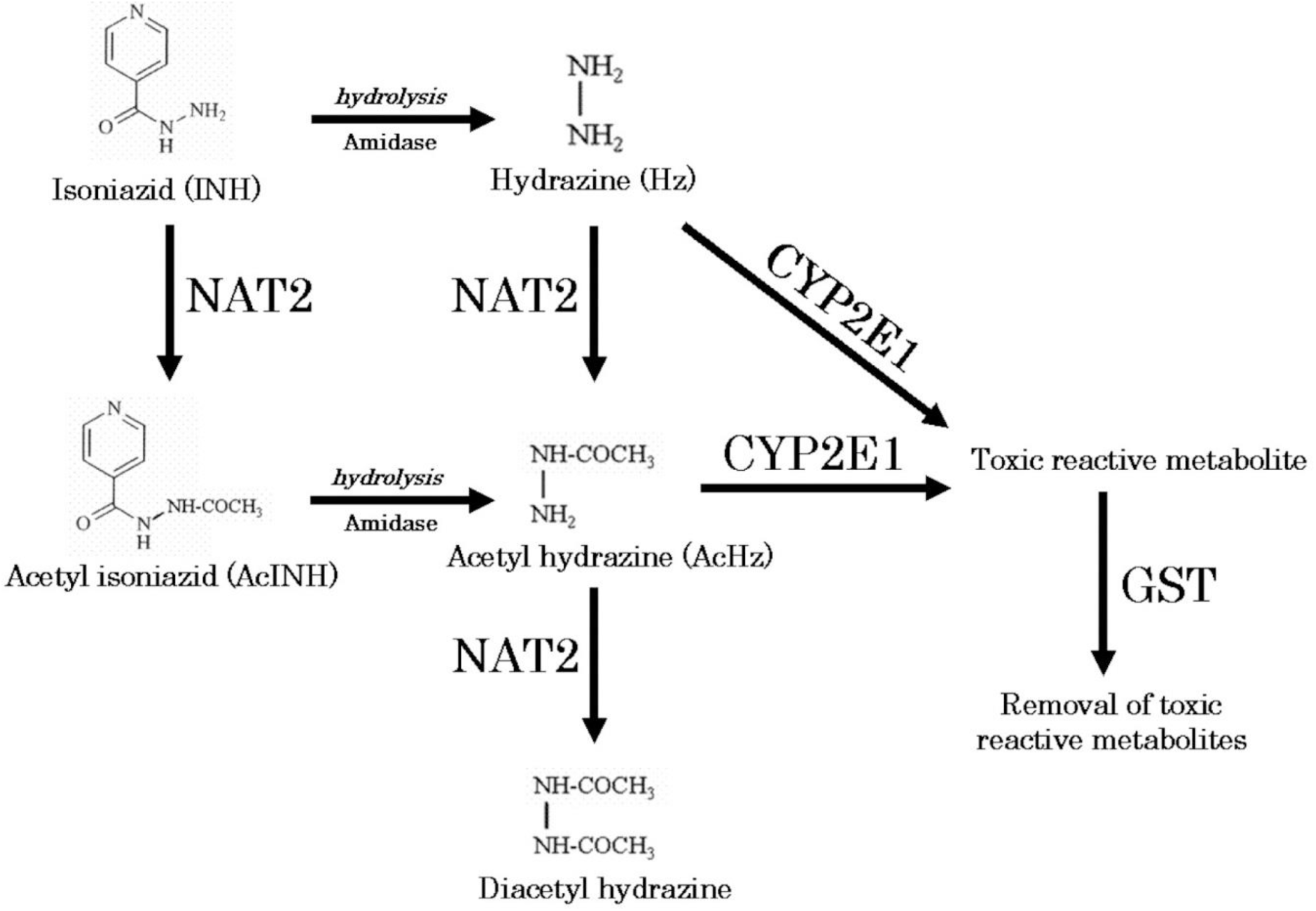
Metabolic pathway of isoniazid^36^. Reproduced with permission

### Physiologically-based pharmacokinetic model of liver tissue exposures of isoniazid and its metabolites

To predict the hepatic tissue exposures of isoniazid, acetylisoniazid, hydrazine, and acetylhydrazine during isoniazid treatment, the whole-body physiologically based pharmacokinetic model with advanced compartmental and transit model in GastroPlus (Ver 9.7) was used (Supplementary Figure 1). All physicochemical properties for those substances were predicted based on chemical structure from a built-in predictor in GastroPlus. Population-dependent physiological parameters, including tissue volumes and perfusion rate in humans, were obtained using the Population Estimates for Age-Related Physiology module in GastroPlus (Supplementary Table 1). After simulating time-versus-concentration profiles of substances in liver and plasma, noncompartmental pharmacokinetic analysis was performed to calculate the AUC_0-24_ ratio between liver and plasma. This ratio was used to predict liver tissue exposures to isoniazid and metabolites, during a 24-hour dosing interval, based on the observed plasma exposures in the clinical study.

### *In vitro* pharmacologic exposures of hepatocytes to isoniazid and its metabolites

The human liver carcinoma cell line, HepG2, is highly differentiated and is often used to screen the cytotoxicity potential of new chemical entities early in drug development^14^. HepG2 cells were cultured in Dulbecco’s modified Eagle’s medium (DMEM) supplemented with 10% fetal bovine serum (FBS), 100 U/mL penicillin, 100 μg/ml streptomycin and 2mM L-glutamine, unless otherwise stated. Cells were continuously incubated at 37°C in a humidified 5% CO2 / 95% air environment and plated in a 12-well plate at 4⨯10^5^ cells/well, over a duration of 24 hours prior to compound exposure. Compound stocks dissolved in water were freshly made each exposure day. Cellular supernatant was replaced with 2 mL of culture media containing 10 µM of compound and left to incubate until 90% confluency was reached. Cell passages were conducted every 3 days under continual exposure to the designated compound for the indicated time period. Cellular supernatant was collected at each passage, clarified, aliquoted, and stored at -80°C for future analysis. Treated cells and supernatants were then examined for markers of cytotoxicity, mitochondrial toxicity, and oxidative DNA damage as described below.

#### Free thiol availability assay

By serving as targets of ROS, thiols provide the cellular defense system against oxidative stress. Glutathione is the principal non-protein thiol in most cells, and high intracellular glutathione concentrations are required to maintain protein thiols in reduced states, which is essential for cellular functions in metabolism, signaling, and detoxification^15^. Cellular glutathione depletion was quantified in a free thiol availability assay^16^. Black 96-well polystyrene clear bottom plates were coated with 50 µg/mL of poly-d-lysine for 1 hour at 37°C. Following multiple washes with PBS, HepG2 cells from the long-term low-exposure culture growth, at the indicated times, were plated at 3.3⨯10^5^ cells per well and left to incubated at 37°C for 12 hours maintain compound exposure. ThiolTracker™ Violet dye (ThermoFisher) was used to asses free thiol availability following the manufacturer’s instructions. Briefly, cells were washed two times with D-PBS +C/M prior to the addition of 20 µM of ThiolTracker. Following a 30 min incubation at 37°C, cells were washed three times with D-PBS +C/M. Fluorescent intensity at 404nm_Ex_/526nm_Em_ was measured on a Synergy H1 Hybrid Multi-Mode Reader (BioTek).

#### Mitochondrial membrane integrity

To assess mitochondrial membrane integrity, black 96-well polystyrene clear bottom plates were coated with 50 µg/mL of poly-d-lysine for 1 hour at 37°C. Following multiple washes with PBS, HepG2 cells from the long-term low-exposure culture growth, at the indicated times, were plated at 3.3×10^5^ cells per well and left to incubated at 37°C for 12 hours maintaining compound exposure. Tetramethylrhodamine, methyl ester (ThermoFisher, Waltham, Massachusetts) was added at 100 nM at 37°C for 30 minutes. Following three washes with PBS, fluorescent intensity at 488nm_Ex_/570nm_Em_ was measured on a Synergy H1 Hybrid Multi-Mode Reader (BioTek).

#### Cellular and mitochondrial toxicity

Mitochondrial toxicity analysis was assayed using the Mitochondrial ToxGlo Assay (Promega, Madison, WI) following the manufacturer’s instructions. Briefly, cells obtained from the long-term low-exposure culture growth, at the indicated times, were grown in a 96-well flat clear bottom black polystyrene plate at 3.3⨯10^5^ cells per well in glucose-free (galactose supplemented) culture media. Compound concentrations (10 µM) were consistently maintained throughout the experiment. Following a 24-hour incubation with compound, cytotoxicity was evaluated by the addition of a fluoregenic peptide substrate, bis-AAF-R110, to the cell supernatant for 30 minutes at 37°C. The bis-AAF-R110 substrate penetrates compromised plasma membranes and is activated via interactions with necrosis-associated proteases. After the cells were allowed to incubate for 30 minutes at 37°C, fluorescence intensity was measured at 485nm_Ex_/525nm_Em_. To evaluate mitochondrial toxicity, the plate was acclimated to room temperature for 5 minutes, at which point the ATP detection reagent was added. Cell lysis mixture was transferred to a 96-well flat solid white bottom plate and luminescence intensity was measured. Assays of fluorescence and luminescence activity were performed with a Synergy H1 Hybrid Multi-Mode Reader (BioTek, Winooski, VT).

#### Comet chip imaging of cellular DNA damage

Cells treated for 1 week with 10 µM of compound were plated onto a CometChip (Trevigen) at 2⨯10^5^ cells/mL in a single cell suspension^17^. Cells were incubated on the chip for 60 min at 37°C. The plate was then gently washed with PBS and coated with 1% low melting agarose. After the agarose was solidified by cooling at 4°C, the plate was washed with pre-chilled non-activated alkaline lysis buffer (2.5 M NaCl, 100nM Na_2_EDTA, 10mM Tris, pH 10). The chip was lysed overnight at 4°C by submerging the chip in the alkaline lysis buffer containing 1% Triton-X. The plate was then washed with PBS and placed in an electrophoresis chamber filled with pre-chilled alkaline electrophoresis buffer (2mM Na_2_ EDTA, 300 mM NaOH). Following a 40 min incubation at 4°C to allow for alkaline unwinding, electrophoresis was carried out at 80 volts for 30 min at 4°C. The chip was then neutralized with two incubations at 4°C in 400 mM Tris, pH 7.5 buffer, and equilibrated in 20 mM Tris, pH 7.4 at 4°C for 20 min. DNA staining was achieved by incubating the chip in 100 mL of 0.2X SYBR Gold overnight at 4°C. Fluorescent images were acquired on an Evos FL Imaging System (ThermoFisher).

#### Immunofluorescence for γ-H2Ax

H2Ax is a core histone protein found in the nucleosome, and the phosphorylation of H2Ax is a marker of DNA damage caused by ROS. Variant histone H2AX (γ-H2AX) is a marker of DNA double-strand breaks^18^. Exposed cells (1 week with 10 µM of compound) were plated in a 12-well cell culture treated plate at 1×10^5^ cells/well. Following 24 hours incubation at 37°C in media containing 10 µM of compound, cells were fixed with a cold 1:1 solution of methanol and acetone for 20 min at -20°C. The cells were subsequently washed with PBS and blocked for 2 hours at room temperature with 1 mL of BlockAid™ Blocking Solution (Thermo Scientific). Each well was stained with a 1:1000 dilution of rabbit anti-phospho-H2AFX (Millipore Sigma, St. Louis, MO) in PBS overnight at 4°C with gently rocking. Following multiple washing steps with PBS, cells were incubated with 5 µg/mL of anti-rabbit Alexa Fluor 647-conjugated antibody (ThermoFisher) for 1 hour at room temperature. Cells were subsequently washed with PBS and stained with 1 µg/mL of Hoechst 33342 (ThermoFisher) for 5 min at room temperature. Fluorescent images were acquired on an Evos FL Imaging System (ThermoFisher).

#### 8-Hydroxy-2’-deoxyguanosine (8-OHdG) in cellular supernatant

Determination of 8-OHdG content in cellular supernatant was assed using an in-house competition ELISA. Clear 96-well plates were coated with 8-Hydroxy-2’-deoxyguanosine (BioVision, Milpitas, CA) overnight at 4°C. Plates were then washed three times with PBS. Cell supernatant in PBS or standard 8-OHdG standard curve dilutions were added to the appropriate wells. Rabbit anti-8-OHdG (Abcam, Cambridge, MA) was subsequently added at a 1:2500 final dilution in PBS and the plate was incubated at room temperature with shaking for 2 hours. Following three washes with PBS, anti-rabbit HRP antibody was added at a 1:5000 dilution and the plate was incubated at room temperature with shaking for 1 hour. Immediately following five rapid PBS washes, 3.7mM o-phenylenediamine in 0.05 M phosphate-citrate buffer containing 0.03% sodium perborate was added and the plate was incubated at room temperature protected from light for 30 minutes. Absorbance at 405 nm was then measured on a Synergy H1 Hybrid Multi-Mode Reader (BioTek).

### Clinical validation of urine oxidative DNA damage in HIV/TB patients

#### Study setting and subjects

We conducted a prospective cohort study of isoniazid pharmacokinetics among HIV/TB patients at 22 public clinics and Princess Marina Hospital in Gaborone, Botswana. The study population has been previously described^19^. Eligibility criteria included citizens of Botswana, HIV infected adults (21 years of age or older) naïve to ART therapy, and newly diagnosed with pulmonary TB initiated on standard first- line TB regimens at weight-based dosing bands recommended in accordance with WHO guidelines. The study visit occurred between 5 and 28 days after initiation of first-line TB therapy that included isoniazid, prior to initiation of ART therapy.

#### Data collection

After an overnight fast, the anti-TB drugs were directly administered. Blood samples (10 mL) were drawn at 0, 0.3, 0.9, 2.2, 4.5, and 8 hours post-dosing, based on optimal sampling theory to estimate isoniazid pharmacokinetic parameters. Plasma isoniazid concentrations were measured with liquid chromatography-tandem mass spectrometry (LC-MS/MS) methods as previously described. The peak plasma concentration (C_max_) was obtained directly from the concentration-versus-time data), model- predicted isoniazid concentrations were used to estimate the area under the concentration-versus-time curve during a 24-hour interval (AUC_0-24_). A spot urine sample (approximately 50 mL) was collected 4 hours following anti-TB drug dosing and frozen at -80C until analysis. We measured urinary concentrations of 8-OHdG, a marker of oxidative DNA damage that is stable in cryopreserved urine samples, normalized to urine creatinine. The DNA Damage Competitive ELISA kit (ThermoFisher Scientific) was performed using a 1:4 dilution of urine-to-assay buffer. The manufacturer’s instructions were followed with one modification to the protocol. Antigen binding was performed overnight at 4°C with shaking in lieu of a 2-hour incubation at room temperature. Each assay was performed in triplicate. Optical absorption was assessed using a Synergy H1 Hybrid Multimode Reader (BioTek). To obtain creatinine concentrations for all urine, the Creatinine Urinary Detection Kit (ThermoFisher Scientific) was performed according to the manufacturer’s instructions using a 1:20 dilution of urine-to-deionized water.

#### Pharmacogenetic genotyping for *NAT2* and *GST* family genes

DNA from whole blood samples was used to perform whole exome sequencing as previously described^19^. Individuals with any combinations of the following NAT-2 alleles: 2*4, 2*11, 2*12 and 2*13, were classified rapid acetylators, while those with both combination of these alleles: 2*5, 2*6, 2*7 and 2*14, were considered slow acetylators, in accordance with existing literature^20^. Patients who possessed one allele from the former group and another from the later were classified as intermediate acetylators. Patients whose single-nucleotide polymorphism (SNP) calls were not available in public databases (http://nat.mbg.duth.gr/ and http://NAT-2pred.rit.albany.edu/) or published literature were designated as ambiguous. We evaluated allele frequencies in SNPs corresponding to GST family genes based on previously identified clinical associations with oxidation/reduction status or isoniazid hepatotoxicity, including *GSTA2, GSTP1*, and *GSTM1*^*21,22*^. Based on a previously published classification scheme^23^, we categorized *GSTA2* haplotypes according to four SNPs (rs2180314, rs6577, rs2234951, rs1803682), as shown in Supplementary Table 3.

### Statistical analysis

Pharmacokinetic analyses were performed in Phoenix 8.0 (Certara USA, Inc., Princeton, NJ), and physiologically based pharmacokinetic modeling was performed GastroPlus v9.7 (SimulationsPlus, Redwood City, CA). Multivariate regression modeling was performed in Stata v13 (College Station, TX: StataCorp LP). Plots were generated in GraphPad Prism version 7.00 for Windows (GraphPad Software, La Jolla, CA, www.graphpad.com). Statistical significance was declared for p-values less than 0.05.

## RESULTS

### Pharmacokinetic exposures to isoniazid and its metabolites in TB patients

We obtained parent-metabolite pharmacokinetic data from 10 isoniazid-treated patients, including 4 patients with latent TB infection and 6 patients with active TB disease. The properties of the HPLC-MS/MS plasma assays for isoniazid and metabolites are included in Supplementary Table 2. The spaghetti plots of pharmacokinetic profiles are shown in Figure 2 for isoniazid (Figure 2a), acetylisoniazid (Figure 2b), hydrazine (Figure 2c), and acetylhydrazine (Figure 2d). For isoniazid, acetylisoniazid, and acetylhydrazine, all concentrations were greater than the lower limit of quantification, corresponding to 1 ng/mL, 10 ng/mL, and 10 ng/mL for isoniazid, acetylisoniazid, and acetylhydrazine, respectively. For hydrazine, 6 of 60 concentrations (10%) were below the lower limit of quantification, corresponding to 2 ng/mL. As expected, plasma AUC_0-24_ exposures for isoniazid and hydrazine were highly correlated (R^2^ = 0.78), consistent with the role of NAT2 enzyme in the metabolism of each compound (Figure 1).

**Figure 2.**
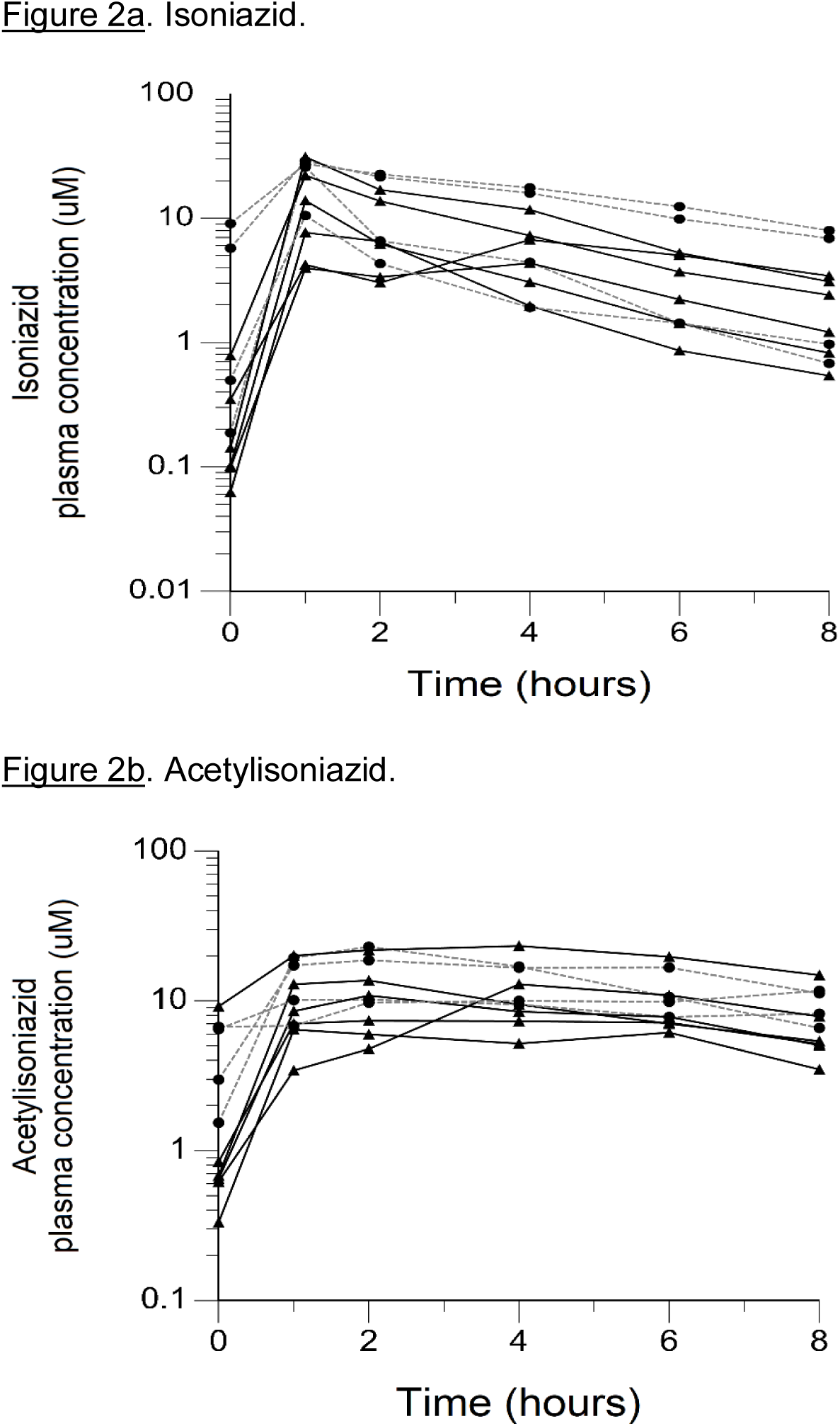
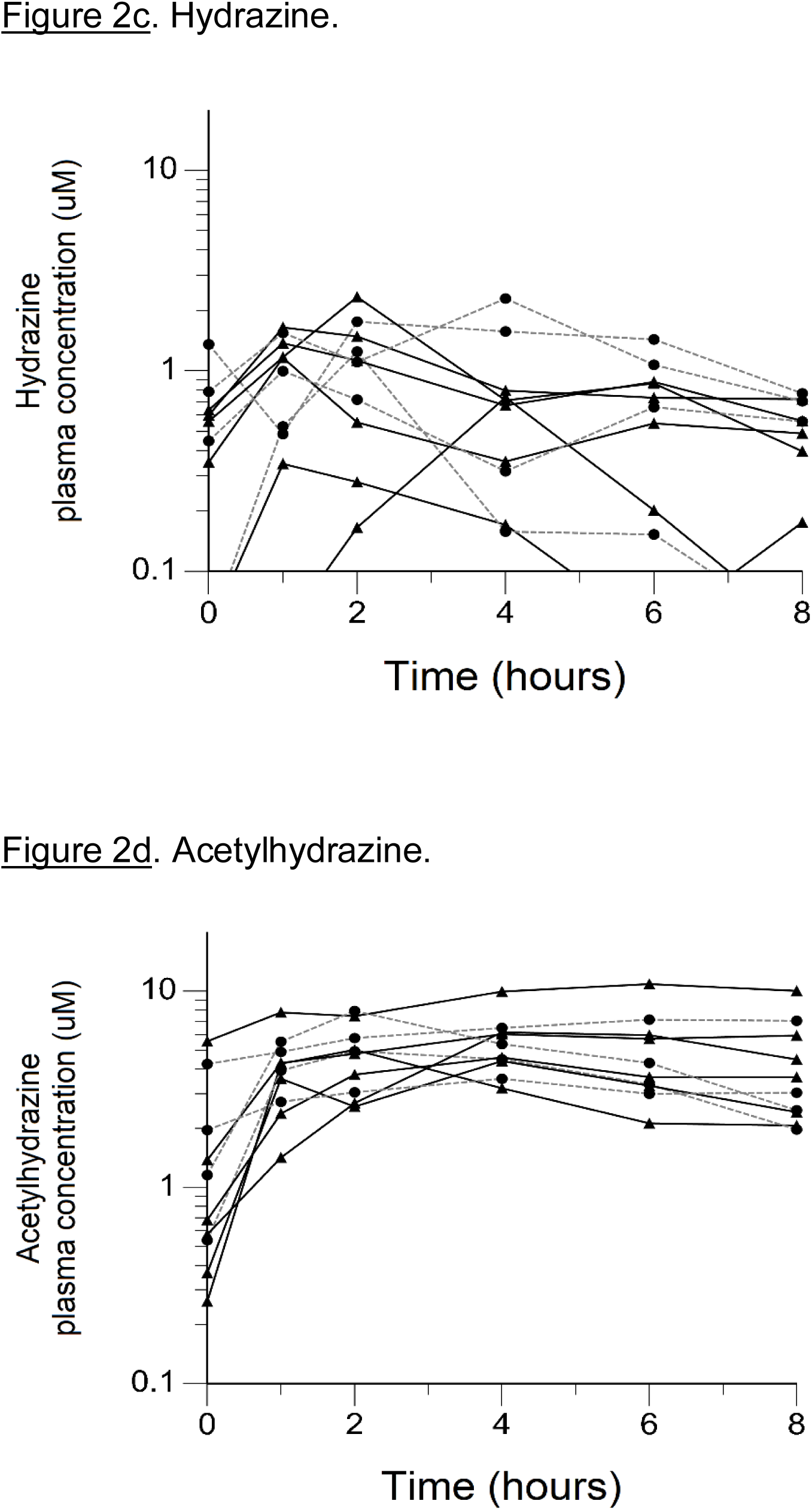
Plasma concentration-time exposures of isoniazid and its metabolites. <u>Figure 2a</u>. Isoniazid. <u>Figure 2b</u>. Acetylisoniazid. <u>Figure 2c</u>. Hydrazine. <u>Figure 2d</u>. Acetylhydrazine. <u>Figure 2e</u>. Correlation of isoniazid and hydrazine AUC_0-24_.

The physiologically based pharmacokinetic model of isoniazid and metabolite distribution into tissue is included in Supplementary Figure 1. Based on this model, we predicted ratios of liver tissue:plasma exposures for isoniazid, acetylisoniazid, hydrazine, and acetylhydrazine as shown in Table 1. The model predicted an 18-fold higher liver tissue exposure for hydrazine compared to plasma, but lower liver tissue exposures for parent compound and the other metabolites compared to plasma. Based on these model-predicted liver tissue exposures, we selected the concentration of 10 uM to study in pre- clinical toxicodynamic models for all compounds, corresponding to an AUC_0-24_ of 240 uM*hr. This molar concentration fell within 1 log-10 for the median exposure to each compound.

**Table 1.**
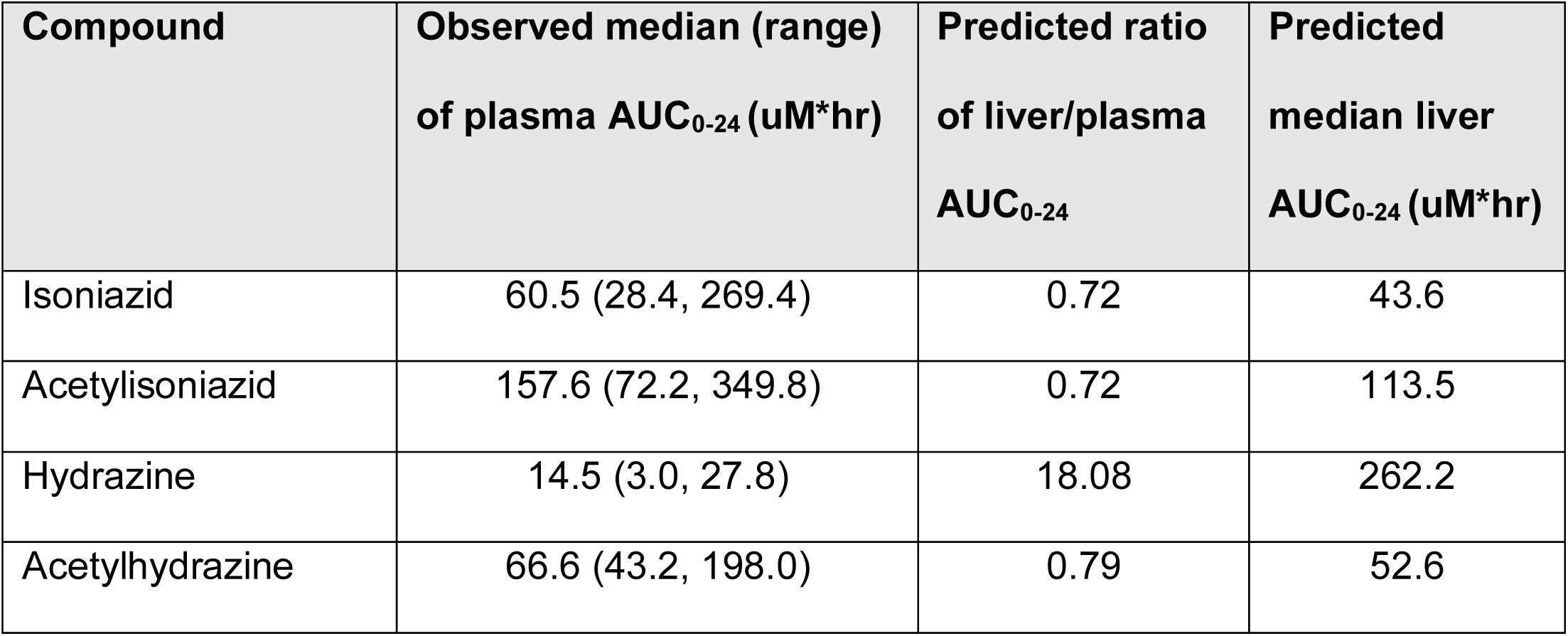
Predicted liver tissue exposures of isoniazid and metabolites.

### Long-term exposures of HepG2 cells treated at clinically relevant concentrations of isoniazid and metabolites

#### Biochemical assays of redox imbalance and mitochondrial injury

There was no appreciable cytotoxicity and of isoniazid or its metabolites (Figure 3a). However, we observed thiol depletion in treated cells over the 28-day exposure period, demonstrating impaired cellular capacity to maintain the oxidation-reduction equilibrium, greatest among hydrazine-treated cells (Figure 3b). TMRM is a cell-permeant dye that accumulates in active mitochondria with intact membrane potential and is depleted with free radical injury to mitochondrial membrane potential. Figure 3c shows that there was TMRM depletion in all treated cells across 28 days of exposure, with the greatest depletion observed for hydrazine. Furthermore, Figure 3d shows that cellular ATP production was impaired at 28 days of treatment across all experiments, again with the greatest impairment observed for hydrazine as compared with the other compounds.

**Figure 3.**
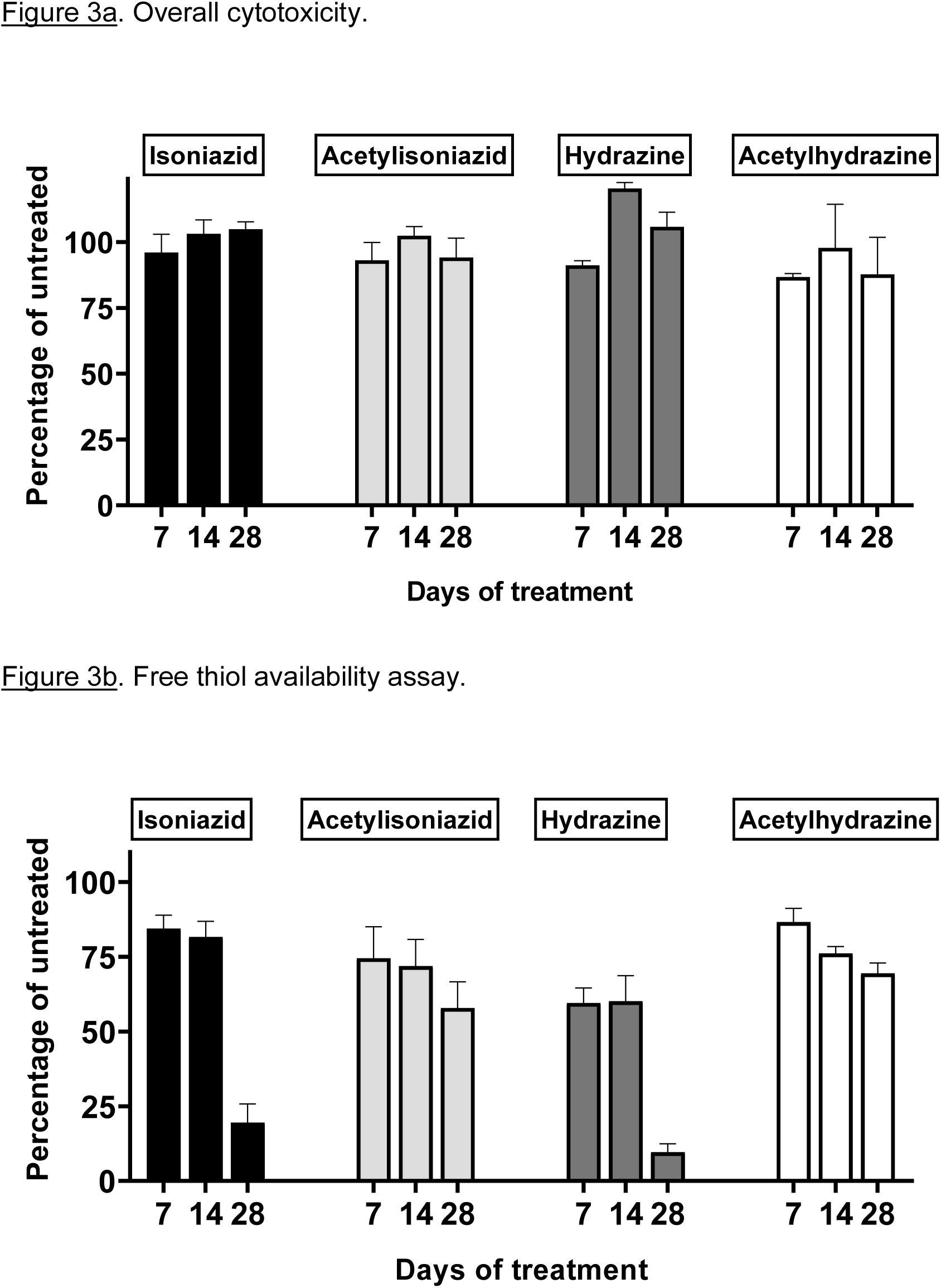
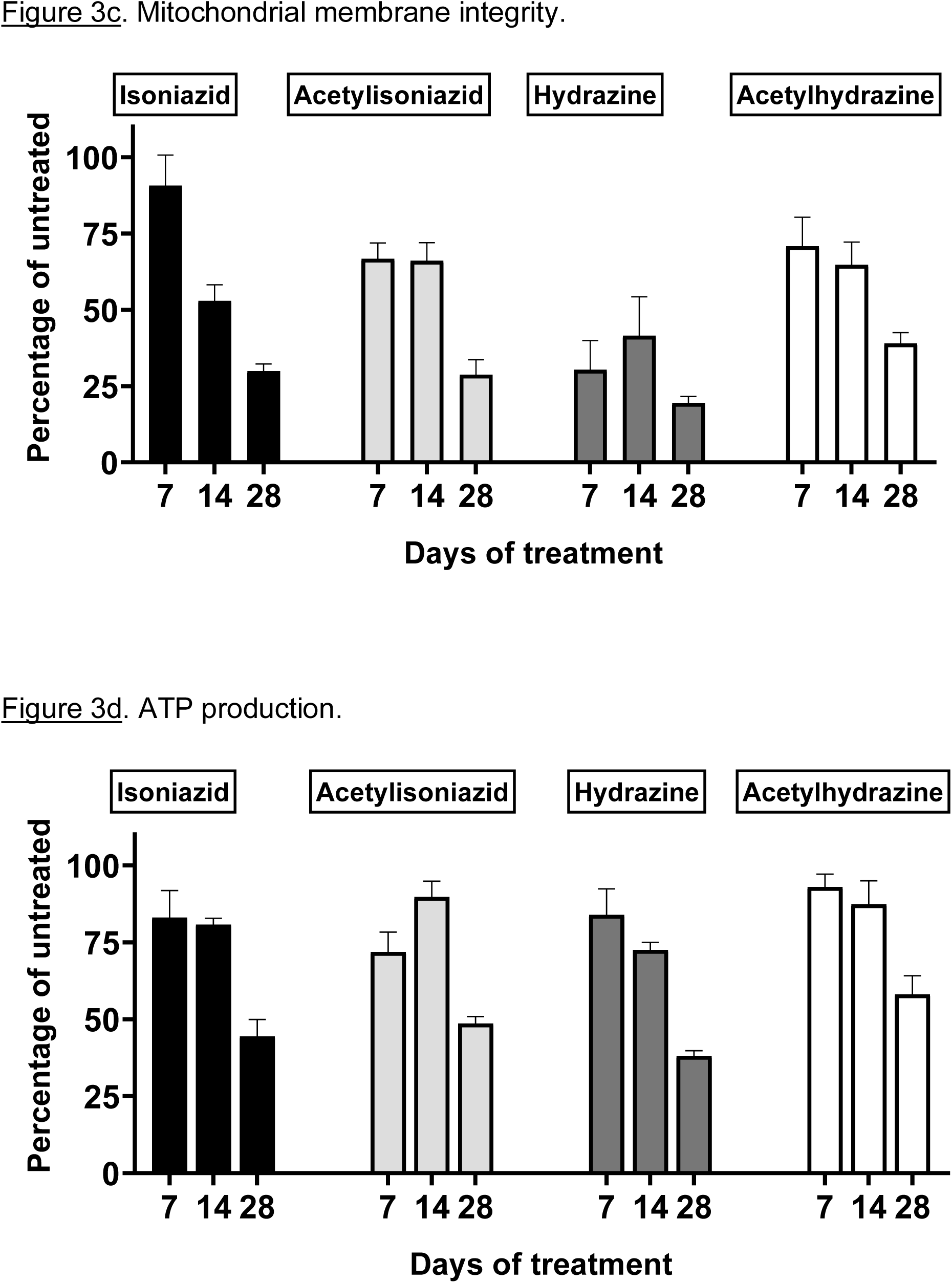
Mitochondrial injury and glutathione depletion in HepG2 cells treated with isoniazid and metabolites over 28 days. <u>Figure 3a</u>. Cytotoxicity. <u>Figure 3b</u>. Free thiol assay. <u>Figure 3c</u>. Mitochondrial membrane integrity. <u>Figure 3d</u>. ATP production.

#### Microscopic and biochemical assays of oxidative DNA damage

We examined levels of oxidative DNA damage in treated HepG2 cells in several ways. First, CometChip microscopy demonstrated greater levels of DNA damage in cells treated for 1 week with hydrazine, acetylhydrazine, and acetylisoniazid, as compared to isoniazid (Figure 4a). Next, we measured γ-H2AX in treated cells, a variant histone which corresponds to the presence of double- stranded DNA breaks. Following a 1-week exposure period, γ-H2AX levels were greatest during treatment with hydrazine, as compared to isoniazid, acetylisoniazid, and acetyhydrazine (Figure 4b). Finally, we examined cellular supernatant levels of 8-OhDG, a measure of oxidative DNA damage that is stable in cryopreserved clinical samples^24^. Figure 4c shows that the highest increase in 8-OhDG levels in cell supernatant was with hydrazine treatment. Given that the 10 µM concentration used in these experiments was less than predicted in liver tissue for hydrazine, but greater than predicted for isoniazid and non-hydrazine metabolites, these findings collectively supported our hypothesis that hydrazine is the primary mediator of redox imbalance in hepatocytes.

**Figure 4.**
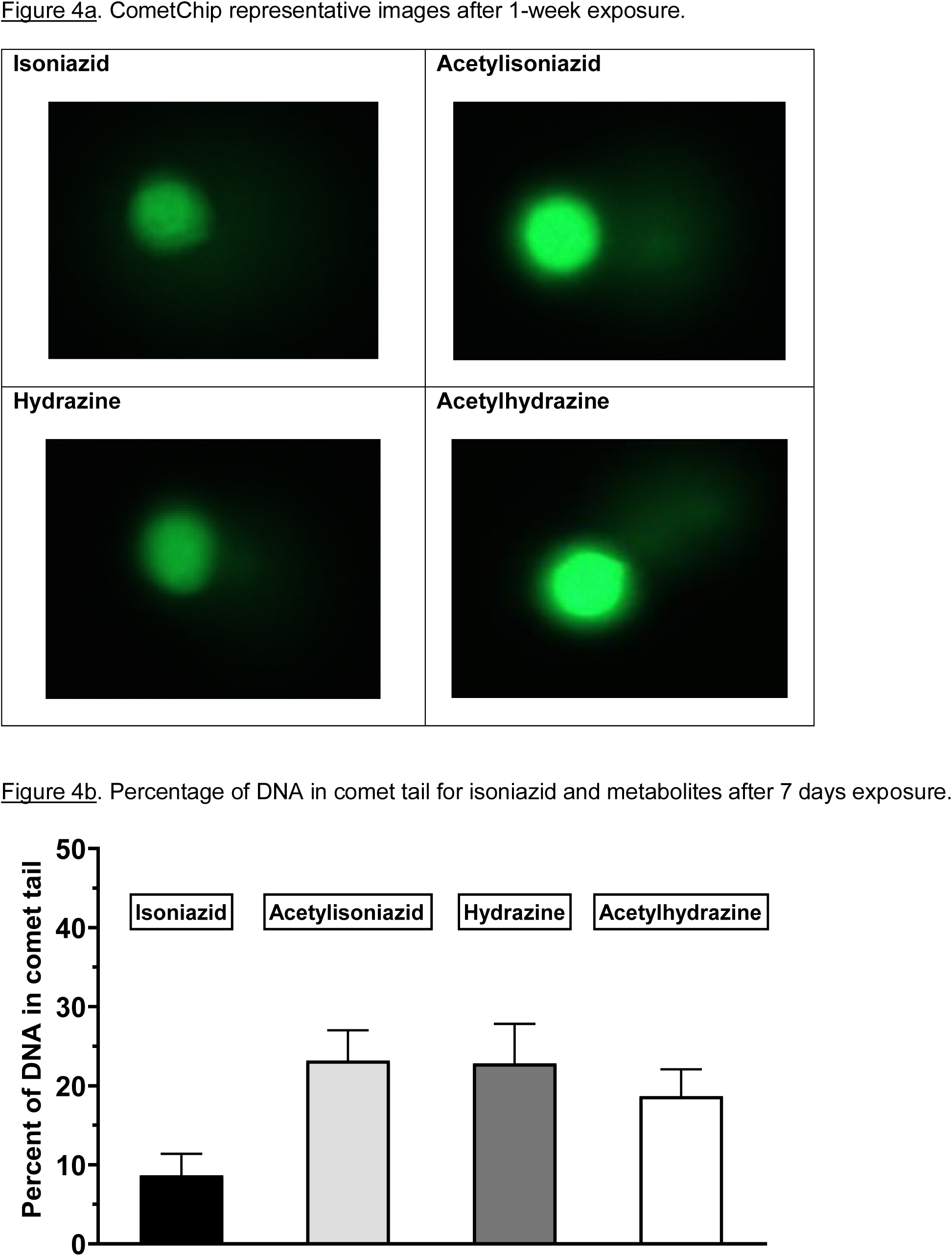
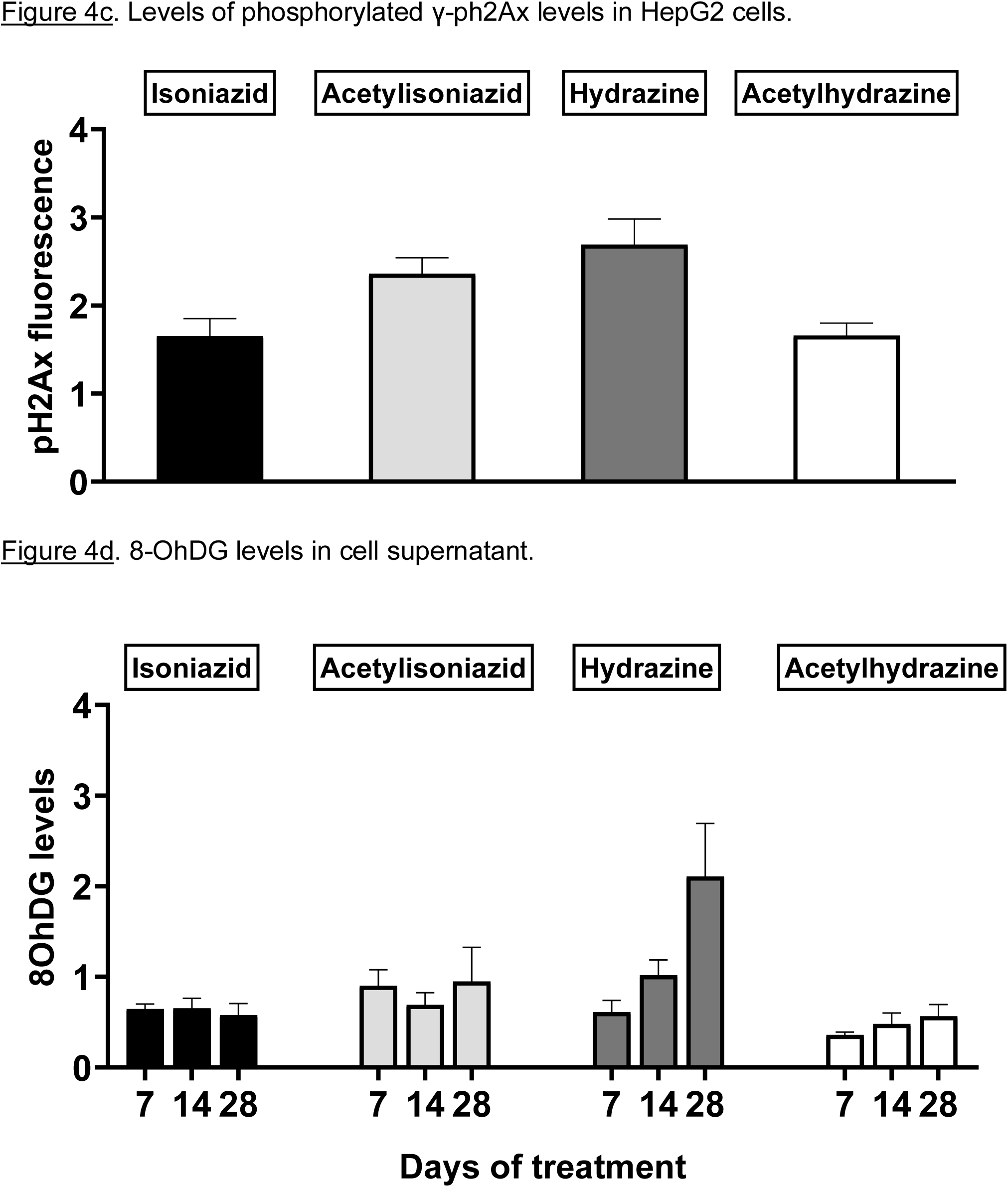
DNA damage markers in HepG2 cells treated with isoniazid and metabolites. <u>Figure 4a</u>. Comet chip microscopy of DNA damage after 7 days exposure. <u>Figure 4b</u>. Percentage of DNA in comettail for isoniazid and metabolites after 7 days exposure. <u>Figure 4c</u>. Levels of phosphorylated gamma- ph2Ax levels in HepG2 cells after 28 days exposure. <u>Figure 4d</u>. 8-OhDG levels in cellular supernatant following 28 days exposure.

### Oxidative DNA damage among isoniazid-treated TB patients

To further test the clinical relevance of these in vitro findings, we collected urine samples obtained in a previously conducted clinical pharmacokinetic study of HIV/TB patients,^19^ to test for 8-OhDG concentrations, a stable marker of oxidative DNA damage, which has been linked to exposures to toxins such as cadmium^25^, chromium, and lead^26^ in other patient cohorts. Demographic, clinical, and immunologic characteristics of study participants are shown in Table 2, which shows that none of the characteristics differed *NAT2* genotype. However, *NAT2* genotype was associated with urine 8-OhDG concentrations, as shown in Figure 5a. In unadjusted analysis, we observed a significant increase in urine oxidative DNA damage among patients with slow NAT2 genotypes, compared to rapid acetylator genotypes (p=0.04). Next, we examined *GSTA2*, which encodes the dominant GST protein in human livers, and demonstrates population variability that corresponds to varying levels of hepatic GST expression^22^. The rs2234951 and rs1803682 variants were not detected in the study cohort, while the SNPs rs6577 and rs2180314 were in complete linkage disequilibrium. Accordingly, all haplotypes were either GSTA2*B or *C under the naming schema proposed by Tetlow *et al*^23^ (Supplementary Table 3). We identified 25 patients with the homozygous *GSTA2*B/*B* haplotype (rs6577/rs6577), 12 patients with the *GSTA2*C/*B* haplotype (rs6577/rs2180314), and 2 patients with the homozygous *GSTA2*C/*C* haplotype (rs2180314/rs2180134). In unadjusted analysis, *GSTA2* genotype (*GSTA2*C/*B* or *GSTA2*C/*C* versus *GSTA2*B/*B*) was associated with urine oxidative DNA damage, as shown in Figure 5b (p=0.03). In contrast, neither the *GSTP1* rs1695 variant, previously identified as a predictor of isoniazid toxicity in a Chinese cohort^21^, nor the *GSTM1* null variant were associated with urine oxidative DNA damage; however the number of patients with the GSTM1 null variant was small (n=3).

**Table 2.** Characteristics of HIV/TB patients, stratified by *NAT2* genotype.

| Characteristic | Slow *NAT2*<br>genotype (N=7) | Intermediate *NAT2*<br>genotype (N=20) | Rapid *NAT2*<br>genotype (N=13) | P-value |
| --- | --- | --- | --- | --- |
| Median age (years) (range) | 30 (24 - 44) | 32 (25 - 48) | 32 (24 - 49) | 0.54 |
| Male sex (%) | 2 (29%) | 11 (55%) | 9 (69%) | 0.22 |
| Median body weight (kg)<br>(range) | 55.0 (44.0 - 67.0) | 54.8 (37.3 - 77.0) | 55.0 (42 - 60.5) | 0.64 |
| Median creatinine clearance<br>(mL/min) (range) | 100.9 (85.3 - 129.6) | 107.2 (68.4 - 141.3) | 98.4 (40.1 - 157.6) | 0.69 |
| Number of current smokers<br>(%) | 0 (0%) | 1 (5%) | 2 (15%) | 0.40 |
| Number with positive AFB<br>sputum smear (%) | 4 (57%) | 15 (79%) | 9 (69%) | 0.53 |
| Median CD4+ T cell count<br>(cells/mL<sup>3</sup>) (range) | 252 (208 - 486) | 238 (36 - 501) | 209 (11 - 735) | 0.69 |
| Median HIV viral load<br>(copies/mL) (range) | 2.5x10<sup>4</sup><br>(1.6x10<sup>3</sup> - 7.0x10<sup>6</sup>) | 1.1x10<sup>5</sup><br>(86 - 5.1x10<sup>6</sup>) | 1.3x10<sup>5</sup><br>(4.3x10<sup>3</sup> - 1.8x10<sup>6</sup>) | 0.72 |

**Figure 5.**
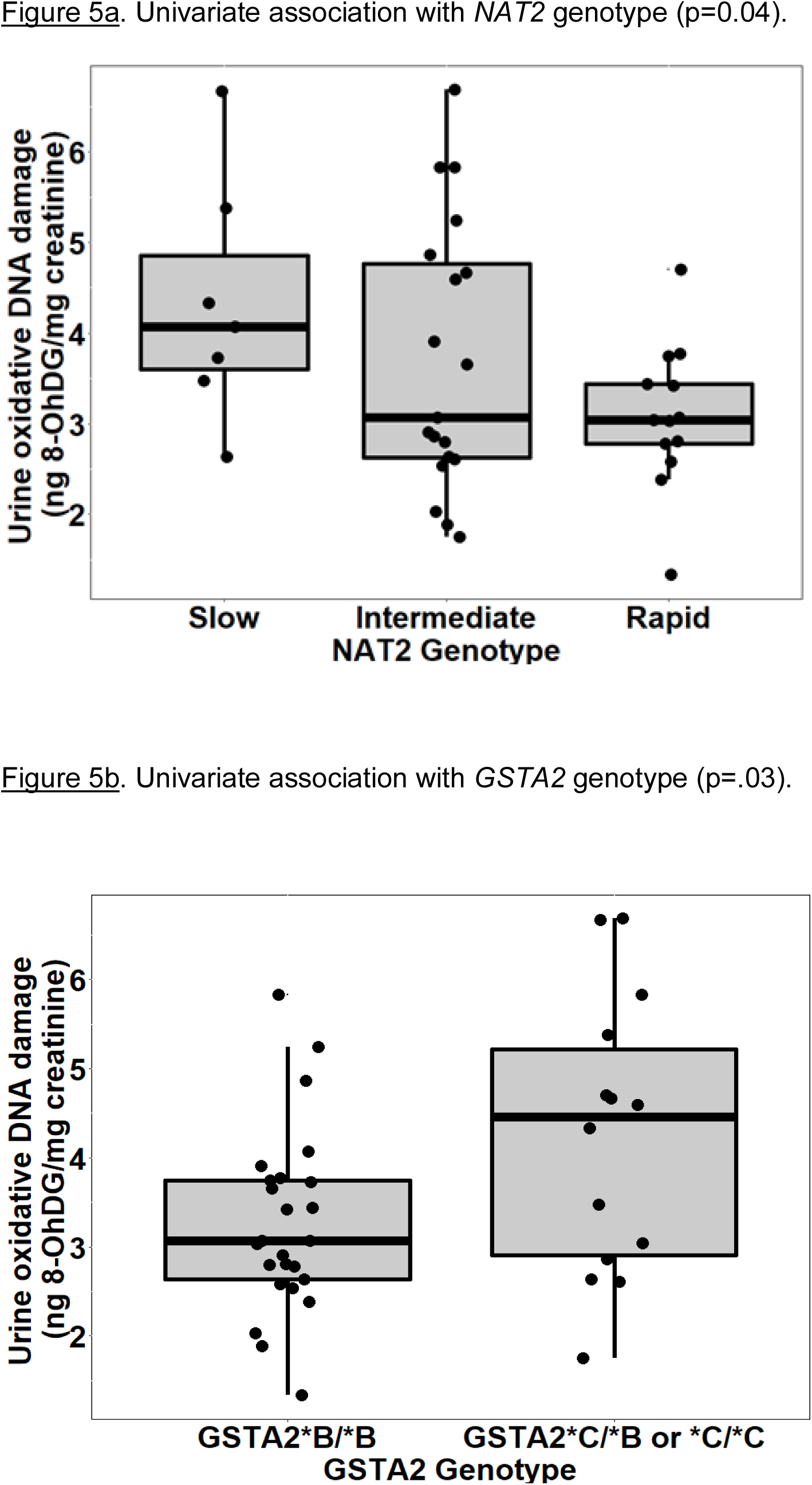
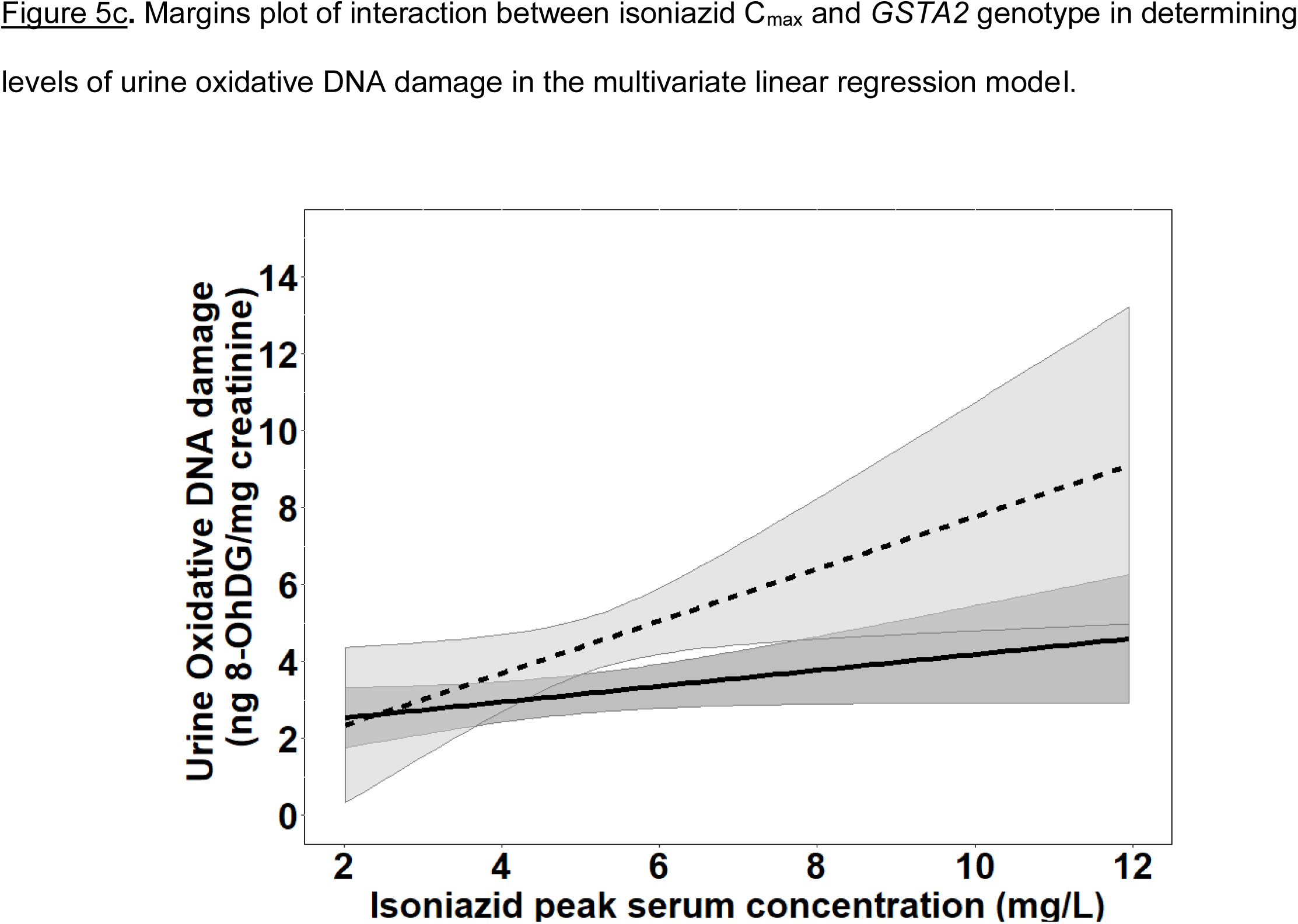
Urine levels of oxidative DNA damage among HIV/TB patients, grouped by NAT2 and GSTA2 genotypes. <u>Figure 5a</u>. *NAT2* genotype and urine oxidative DNA damage. <u>Figure 5b</u>. *GSTA2* genotype and urine oxidative DNA damage. <u>Figure 5c</u>. Margins plot of interaction between isoniazid C_max_ and *GSTA2* genotype. Each line represents marginal predicted levels of oxidative DNA damage for varying isoniazid C_max_, according to *GSTA2* genotype, and the shaded areas represent 95% confidence intervals. Solid line: *GSTA2*B/*B*; dashed line: *GSTA2*C/*B* or *\*C*.

Next, we examined the relationship between isoniazid pharmacokinetic exposures and urine oxidative DNA damage in a multivariate linear regression model. Based on our *a priori* criteria for model inclusion (greater than 20% change in the regression coefficient when the confounder was included in the model), the final multiple linear regression model included the confounding effects of body weight and renal function on the association between isoniazid C_max_ and urine oxidative DNA damage (Table 3). Although not a significant confounder of this relationship, we included the effects of *GSTA2* genotype (*B/*B vs *C/*B or *C/*C) based on its independent association with urine levels of oxidative DNA damage in unadjusted analysis. The corresponding increase in adjusted R-squared with sequential variable inclusion, along with the decline in Akaike information criteria (AIC) score, provided statistical support for choosing model as variables were added. Variance inflation factors for the variables in the final model were 1.31, 2.38, 1.78, and 1.39 for isoniazid C_max_, body weight, creatinine clearance, and *GSTA2* genotype, respectively, supporting the absence of significant co-linearity among model predictors.

**Table 3.** Linear regression model of predictors of urine oxidative DNA damage among isoniazid-treated HIV/TB patients.

| Characteristic | Unadjusted regression coefficient (95% CI) | P-value | Adjusted regression coefficient (95% CI) | P-value |
| --- | --- | --- | --- | --- |
| Age (years) | -0.013 (-0.067, 0.041) | 0.63 |  |  |
| Sex |  | 0.36 |  |  |
| Male (n=) | Reference |  |  |  |
| Female (n=) | 0.391 (-0.464, 1.247) |  |  |  |
| Body weight (kg) | -0.004 (-0.055, 0.047) | 0.88 | -0.094 (-0.164, -0.026) | 0.01 |
| Creatinine clearance (mL/hr) | 0.010 (-0.011, 0.031) | 0.35 | 0.020 (-0.005, 0.045) | 0.11 |
| Positive AFB sputum smear | -0.449 (-1.396, 0.498) | 0.34 |  |  |
| Days of TB treatment | 0.004 (-0.029, 0.038) | 0.80 |  |  |
| CD4+ T cell count (cells/mL<sup>3</sup>) | 0.001 (-0.002, 0.003) | 0.48 |  |  |
| Log HIV viral load | -0.009 (-0.193, 0.175) | 0.92 |  |  |
| CRP | 0.014 (-0.012, 0.040) | 0.28 |  |  |
| Log IL-6 | -0.001 (-0.009, 0.008) | 0.85 |  |  |
| **Isoniazid pharmacokinetic exposures** |  |  |  |  |
| Isoniazid C<sub>max</sub> (mg/L) | 0.153 (-0.060, 0.366) | 0.15 | 0.241 (0.020, 0.462) | 0.03 |
| Isoniazid log AUC<sub>0-24</sub> | 0.474 (-0.272, 1.221) | 0.21 |  |  |
| **Pharmacogenetic variability** |  |  |  |  |
| NAT2 genotype |  | 0.12 |  |  |
| Rapid (n=13) | Reference |  |  |  |
| Intermediate (n=20) | 0.616 (-0.311, 1.544) |  |  |  |
| Slow (n=7) | 1.241 (0.033, 2.449) |  |  |  |
| *GSTA2* genotype |  |  |  |  |
| *GSTA2* *B/*B (n=25) | Reference |  |  |  |
| *GSTA2* *B/*C (n=12) or<br>*GSTA2* *C/*C (n=2) | 0.971 (0.131, 1.810) | 0.03 | 1.259 (0.321, 2.198) | 0.02 |
| *GSTP1* SNP rs1695 |  | 0.34 |  |  |
| WT/WT (n=5) | Reference |  |  |  |
| WT/Var (n=26) | 0.848 (-0.359, 2.054) |  |  |  |
| Var/Var (n=8) | 0.469 (-0.965, 1.902) |  |  |  |
| *GSTM1* null genotype (n=3) | -0.819 (-2.415, 0.777) | 0.31 |  |  |

Figure 5c is a marginal regression plot of isoniazid C_max_ and urine oxidative DNA damage, with effect modification by *GSTA2* genotype. Although the graphical relationship suggests that *GSTA2* genotype acts both as an independent predictor as well as an effect modifier of the relationship between isoniazid C_max_ and urine oxidative DNA damage, the difference in slopes did not reach the *a priori* threshold for statistical significance (p=0.12 for interaction term).

## DISCUSSION

Despite being a cornerstone of TB therapy for 60 years, there is considerable uncertainty regarding the mechanism for isoniazid hepatotoxicity, and the host factors that place TB patients at greatest risk. We first characterized plasma exposures to isoniazid and its metabolites (acetylisoniazid, hydrazine, and acetylhydrazine) in a clinical pharmacokinetic study, demonstrating that isoniazid and hydrazine plasma exposures are highly correlated. Based on these clinical data, we predicted liver tissue exposures using physiologically based pharmacokinetic modeling, observing that hydrazine concentrates in liver relative to plasma. We then developed a long-term, low-exposure experimental set-up in HepG2 cells in order to replicate parent-metabolite pharmacologic exposures in the liver during the course of isoniazid treatment. We found that the greatest levels of mitochondrial injury and oxidative DNA damage occurred with hydrazine treatment at equimolar concentrations. Finally, In a clinical translation of this observation, we observed that peak isoniazid serum concentrations and *GSTA2* genotype were independently associated with increasing urine levels of oxidative DNA damage.

These findings link pharmacokinetic exposures to hydrazine, an isoniazid metabolite, with downstream effects on redox imbalance, adding to prior clinical studies of predictors of isoniazid hepatotoxicity. Slow acetylators of isoniazid, as defined by *NAT2* genotype, have an increased risk of hepatotoxicity compared to intermediate or rapid acetylators, possessing one or two rapid *NAT2* alleles, respectively^27^. In a prior clinical study, peak serum isoniazid concentrations were directly correlated with elevations in liver transaminases, a marker of hepatic inflammation^28^. Future work should identify whether early markers of oxidative stress among isoniazid-treated patients, such as the urine measure of DNA damage identified in this clinical study, are predictive of subsequent hepatotoxicity events. For example, a cross-sectional study among isoniazid-treated TB patients in India demonstrated an association between hepatotoxicity events and co-incident measurements of plasma oxidative stress markers^12^.

The *GSTA2* gene encodes a protein sub-unit of α-GST, the predominant GST enzyme expressed in human liver tissue^23^. Previously published data demonstrated decreased α-GST expression in liver tissue among individuals either heterozygous or homozygous for the *GSTA2*\*C allele, suggesting greater vulnerability to oxidative hepatic injury^22^. Consistent with this observation, we found that the *GSTA2*C* haplotype, either heterozygous or homozygous, was an independent predictor of urine oxidative DNA damage among HIV/TB patients. In contrast, we observed no relationship between urine oxidative DNA damage and the rs1695 allele in *GSTP1*, previously linked to anti-TB drug toxicity among Chinese patients^21^, or the null mutant GSTM1.

The involvement of glutathione pathways in the development of isoniazid hepatotoxicity could identify both preventive and therapeutic strategies. Hydrazine cellular effects include inhibition of complex II of the electron transport chain, leading to accumulation of ROS that overwhelms cellular glutathione stores, and directly damaging to DNA through free radical injury^29^. N-acetylcysteine, clinically used to counter acetaminophen toxicity, has also been used for glutathione deficiency in a wide range of conditions^30^. Identification of intermediary signals on the pathway towards clinical hepatotoxicity events, such as oxidative DNA damage, would also improve efficiency of future clinical trial design of novel TB drug therapies, as isoniazid may be included in these investigational drug combinations^3^.

Aside from hepatotoxicity, oxidative DNA damage during isoniazid treatment may have additional downstream consequences. DNA damage signals induce the nuclear activity of poly(ADP-ribose) polymerase-1 (PARP-1), which is involved in DNA repair activities through multiple pathways^31^. Chronic PARP-1 activation competes for intracellular NAD+, resulting in impaired mitochondrial ATP production and cellular apoptosis^32^. Thus, when considering the clinical implications of increased markers of oxidative DNA damage, additional effects aside from hepatotoxicity events should be considered. For example, isoniazid treatment has been linked to increased markers of cellular apoptosis in immune cells, and direct effects of isoniazid on the patient’s immune-metabolism has been suggested. Intriguingly, isoniazid treatment in mice induced apoptosis markers in CD4+ T cells^33^. Future clinical studies should directly compare isoniazid and metabolite pharmacokinetic exposures with toxicodynamic markers of redox imbalance, including urine markers of oxidative DNA damage.

This clinical and translational study of redox imbalance related to isoniazid and its metabolites had several important limitations. Rather than directly measure concentrations in liver tissue, we used physiologically based pharmacokinetic models to estimate liver tissue exposures for each compound. In the clinical validation study, the metabolite concentrations were not measured, and thus we cannot directly evaluate the relationship between metabolite exposures (such as hydrazine) and oxidative stress in this clinical cohort. Based on our pre-clinical observations, we hypothesize that hydrazine exposures, rather than isoniazid exposures, would demonstrate the greatest association with urine oxidative DNA damage among isoniazid-treated TB patients. Furthermore, we did not perform prospective serial monitoring of hepatic transaminases in the clinical cohort, and the significance of increased urine oxidative stress markers during isoniazid treatment remains uncertain. Prospective studies of oxidative stress markers, particularly the urine measure of oxidative DNA damage identified in this study, will be essential to unravel the link between isoniazid and metabolite exposures, hepatocellular oxidative stress, and clinical hepatotoxicity events.

This study had several strengths. Unlike many earlier studies, we examined physiologically relevant isoniazid and metabolite concentrations, supported by a parent-metabolite clinical pharmacokinetic study and physiologically-based pharmacokinetic modeling^34^. We *in vitro* experiments to provide a mechanistic link between physiologic exposures to these compounds and cellular signals for redox imbalance and oxidative DNA damage. In this approach, we identified a marker of oxidative DNA damage, 8-OHdG, which was strongly induced by hydrazine exposures in the cell model, which we then related to isoniazid pharmacokinetic exposures and *GSTA2* genotype in a clinical study. The identification of a urine-based biomarker of oxidative stress related to pharmacokinetic exposures and/or patient genotype supports non-invasive sampling strategies in future prospective studies of isoniazid toxicity. The feasibility of this future work is enhanced by the potential for point-of-care detection of 8-OHdG in paper-based devices^35^.

In summary, the hydrazine metabolite of isoniazid was the strongest inducer of redox imbalance and downstream oxidative DNA damage in HepG2 cells over long-term exposures to clinically relevant concentrations. In a study of isoniazid-treated TB patients, peak serum isoniazid concentrations and *GSTA2* genotype were independently associated with urine levels of 8-OHdG, a marker of oxidative DNA damage. Future clinical studies should evaluate urine 8-OHdG as an early predictor of clinical hepatotoxicity during isoniazid treatment.

## Data Availability

Data is available with appropriate regulatory approvals.

## Ethics statement

The institutional review boards of the University of Pennsylvania, Botswana Ministry of Health, and the Princess Marina Hospital approved the clinical study. All participants provided written informed consent.

## FIGURE LEGENDS

**Supplementary Figure 1.**
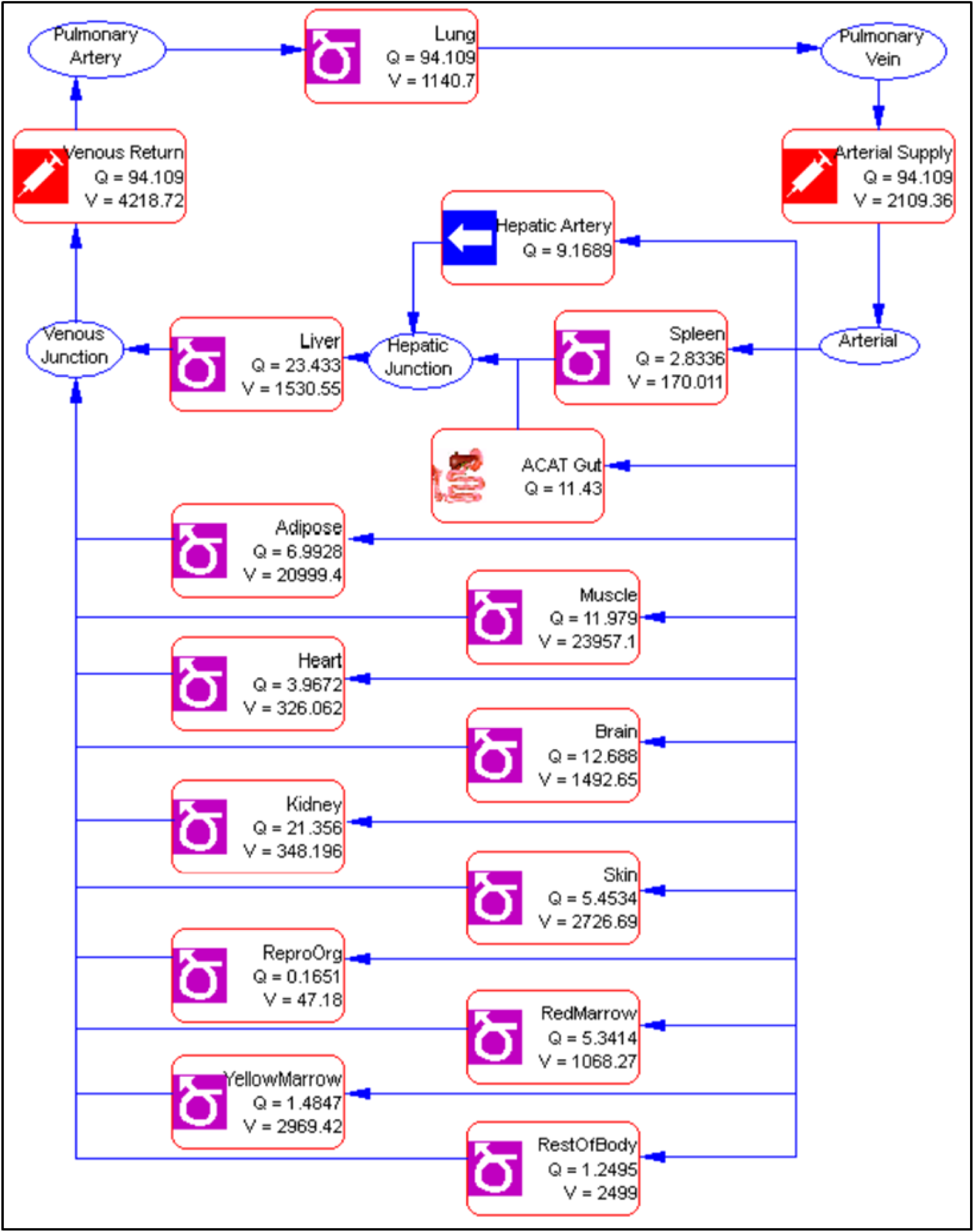
Physiologically-based pharmacokinetic model of isoniazid and metabolites.

**Supplementary Table 1.** Parameters used in the physiologically-based pharmacokinetic model of isoniazid and metabolites.

| Tissue Compartment | Volume (L) | Simulated $K_p$ | | | |
| --- | --- | --- | --- | --- | --- |
|  |  | Isoniazid | Acetylisoniazid | Hydrazine | Acetylhydrazine |
| Lung | 1.14 | 0.72 | 0.70 | 14.31 | 0.79 |
| Arterial supply | 2.11 | 0 | 0 | 0.00 | 0.00 |
| Venous return | 4.22 | 0 | 0 | 0.00 | 0.00 |
| Adipose | 21.00 | 0.17 | 0.17 | 1.55 | 0.18 |
| Muscle | 23.96 | 0.66 | 0.64 | 6.48 | 0.73 |
| Liver | 1.53 | 0.67 | 0.65 | 16.54 | 0.74 |
| Spleen | 0.17 | 0.70 | 0.67 | 11.96 | 0.77 |
| Heart | 0.33 | 0.71 | 0.69 | 8.71 | 0.78 |
| Brain | 1.49 | 0.70 | 0.69 | 2.68 | 0.77 |
| Kidney | 0.35 | 0.70 | 0.67 | 18.06 | 0.76 |
| Skin | 2.73 | 0.66 | 0.65 | 5.43 | 0.71 |
| Reproductive organs | 0.05 | 0.70 | 0.68 | 18.06 | 0.77 |
| Red marrow | 1.07 | 0.41 | 0.41 | 3.00 | 0.43 |
| Yellow marrow | 2.97 | 0.17 | 0.17 | 1.55 | 0.18 |
| Rest of body | 2.50 | 0.70 | 0.68 | 11.96 | 0.77 |

**Supplementary Table 2.**
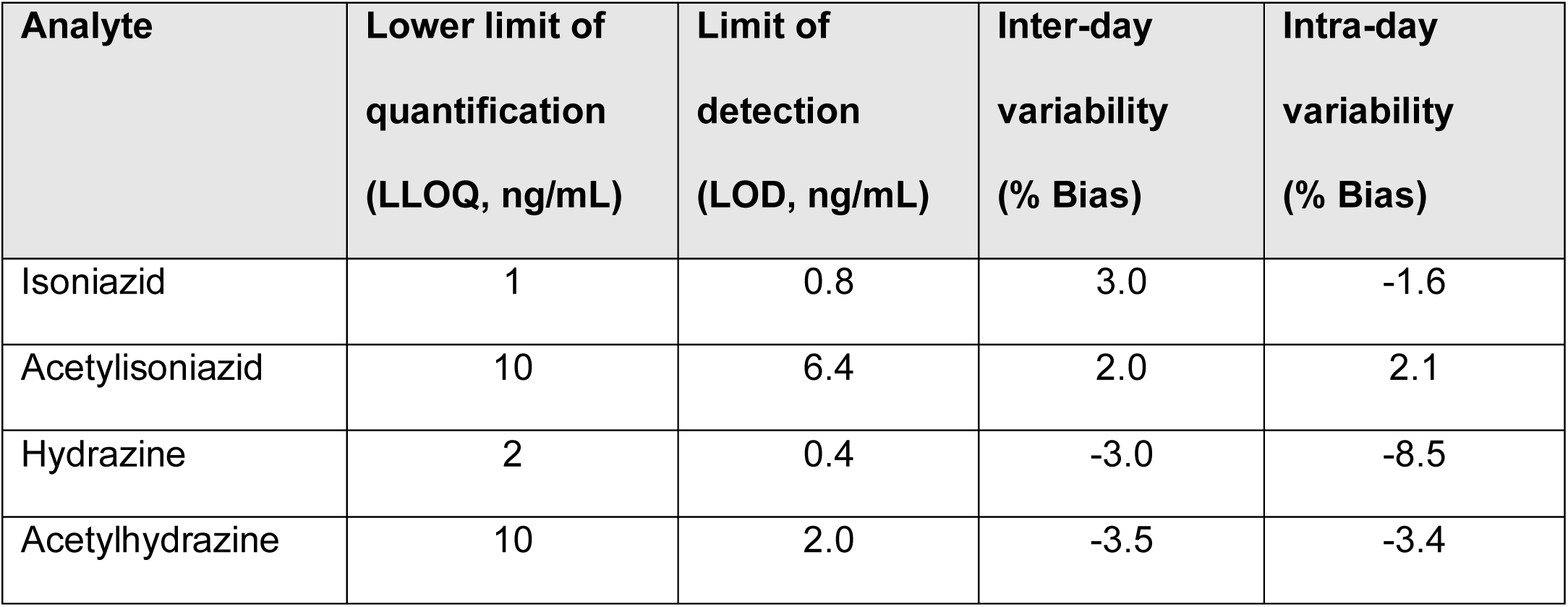
Properties of HPLC-MS/MS plasma assays for isoniazid and metabolites.

**Supplementary Table 3.**
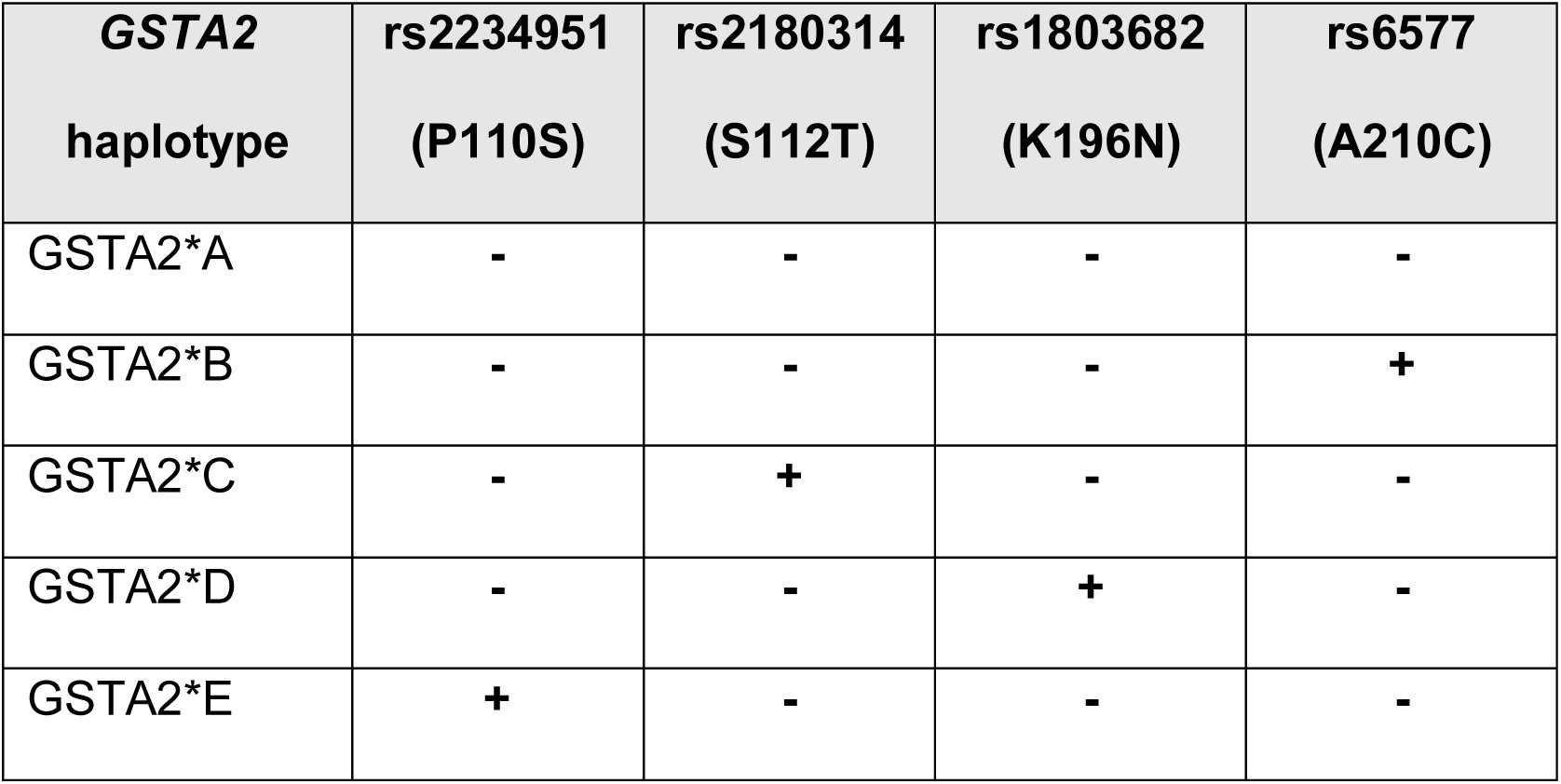
GSTA2 haplotype designations based on SNPs, according to naming schema proposed by Tetlow *et al*. The rs2234951 and rs1803682 variants were not detected in the patient cohort; rs6577 and rs2180314 were in complete linkage disequilibrium.

